# Low-flow in aortic valve stenosis patients with reduced ejection fraction does not depend on left ventricular function

**DOI:** 10.1101/2023.10.19.23297288

**Authors:** Svante Gersch, Torben Lange, Bo Eric Beuthner, Manar Elkenani, Niels Paul, Moritz Schnelle, Elisabeth Zeisberg, Miriam Puls, Gerd Hasenfuß, Andreas Schuster, Karl Toischer

## Abstract

**Background:** Patients with severe aortic stenosis (AS) and reduced left ventricular ejection fraction (LVEF) can be distinguished into high-(HG) and low-gradient (LG) subgroups. However, less is known about their characteristics and underlying (pathophysiological) hemodynamic mechanisms.

**Methods:** 98 AS patients with reduced LVEF were included. Subgroup characteristics were analyzed by a multimodal approach using clinical and histological data, next-generation sequencing (NGS) and applying echocardiography as well as cardiovascular magnetic resonance (CMR) imaging. Biopsy samples were analyzed with respect to fibrosis and mRNA expression profiles.

**Results:** 40 patients were classified as HG-AS and 58 patients as LG-AS. Severity of AS was comparable between the subgroups. Comparison of both subgroups revealed no differences in LVEF (p=0.1), LV mass (p=0.6) or end-diastolic LV diameter (p=0.12). Neither histological (HG: 23.2% vs. LG: 25.6%, p=0.73) and circulating biomarker-based assessment (HG: 2.6 ± 2.2 % vs. LG: 3.2 ± 3.1 %; p= 0.46) of myocardial fibrosis nor global gene expression patterns differed between subgroups. Mitral regurgitation (MR), atrial fibrillation (AF) and impaired right ventricular function (MR: HG: 8% vs. LG: 24%; p<0.001; AF: HG: 30% vs. LG: 51.7%; p=0.03; RVSVi: HG 36.7 vs. LG 31.1 ml/m2, p=0.045; TAPSE: HG 20.2 vs. LG 17.3 mm, p=0.002) were more frequent in LG-AS patients compared to HG-AS. These pathologies could explain the higher mortality of LG vs. HG-AS patients.

**Conclusion:** In patients with low-flow severe aortic stenosis, low transaortic gradient and cardiac output are not primarily due to LV dysfunction or global changes in gene expression, but may be attributed to other additional cardiac pathologies like mitral regurgitation, atrial fibrillation or right ventricular dysfunction. These factors should also be considered during planning of aortic valve replacement.

## Introduction

Aortic stenosis (AS) is the most prevalent valve disease in the ageing society.[1,2] Severe AS occurs in 3-5% of individuals over 75 years and is associated with a high burden of morbidity and mortality.[3] AS patients with reduced left ventricular ejection fraction (LVEF) can be subdivided in two groups: 1) AS with reduced LVEF and high transaortic gradient (Vmax >4.0m/s or PGmean >40 mmHg; HG-AS) and 2) AS with reduced LVEF and low gradient (Vmax <4.0m/s or PGmean <40 mmHg; LG-AS).

With evolving evidence, transcatheter aortic valve implantation (TAVI) has become an established treatment method of severe AS during the past decade. While current guideline clearly recommend AVR in symptomatic patients with HG-AS irrespectively from LVEF, diagnosis and treatment of LG-AS is challenging and an extended multimodal diagnostic approach is suggested in order to achieve diagnostic certainty.[4] Importantly, in a previous study, increased myocardial fibrosis (MF) in these subgroups was shown to be associated with poor outcome.[5] Moreover, LG-AS has been demonstrated to be associated with worse prognosis compared to HG-AS even after aortic valve replacement.[6,7] However, data on HG- and LG-AS phenotypes are scarce and potential underlying hemodynamic mechanisms remain unclear.

Therefore, the aim of this study was to characterize and compare patients with HG-AS and LG-AS in a multimodal approach using clinical data, echocardiography, cardiac magnetic resonance imaging, histology and next-generation-sequencing (NGS) for a more in-depth understanding of these heterogenous AS subgroups.

## Methods

Data for this study were gathered at the University Medical Center Göttingen. Patients were classified into a high-gradient and a low-gradient group according to their transvalvular gradient in echocardiography. According to current guideline recommendations HG-AS was defined as a transvalvular gradient >40 mmHg or a peak aortic jet velocity Vmax > 4m/s and an aortic valve area (AVA) < 1 cm^2^ and LG-AS with a gradient < 40 mmHg and an AVA < 1 cm^2^. In LG-AS group a pseudo stenosis was ruled out either by stress test and/or calcium scoring in computed tomography. On admission, transthoracic echocardiography was performed, a 6-minute walk test, Minnesota Living with Heart failure Quality of life questionnaire (MLHFQ), New York Heart Association (NYHA) status, and N-terminal pro-brain natriuretic peptide (NT-proBNP) levels were measured. Decision for TAVI was based on an interdisciplinary heart team decision according to current guideline recommendations. Additionally, PICP and the CITP:MMP1 ratio, which serves as an indicator of irreversible myocardial fibrosis through collagen cross-linking, was analyzed. A low CITP:MMP1 ratio indicates a presence of irreversible collagen deposition, while a high ratio suggests the possibility of reversible collagen formation. This analysis was conducted using an ELISA-based assay for CITP (Orion Diagnostica, Espoo, Finland) and an alphaLISA method to quantify total serum MMP-1 levels (PerkinElmer, Waltham, Massachusetts, USA) as previously described.[8]

The study complied with the Principles of Helsinki and was approved by the local Ethics Committee Göttingen (Ethic committee Göttingen: 22/4/11). All patients gave written informed before study participation.

### Echocardiography

As previously described[5], all echocardiographies were performed pre-TAVI and were obtained as recommended.[9] Transvalvular pressure gradient was measured by continuity equation and stroke volume (SV) was computed with pulse wave Doppler signal from apical five-chamber view (Figure 1). SV was then indexed to body surface area (BSA).

**Figure 1:**
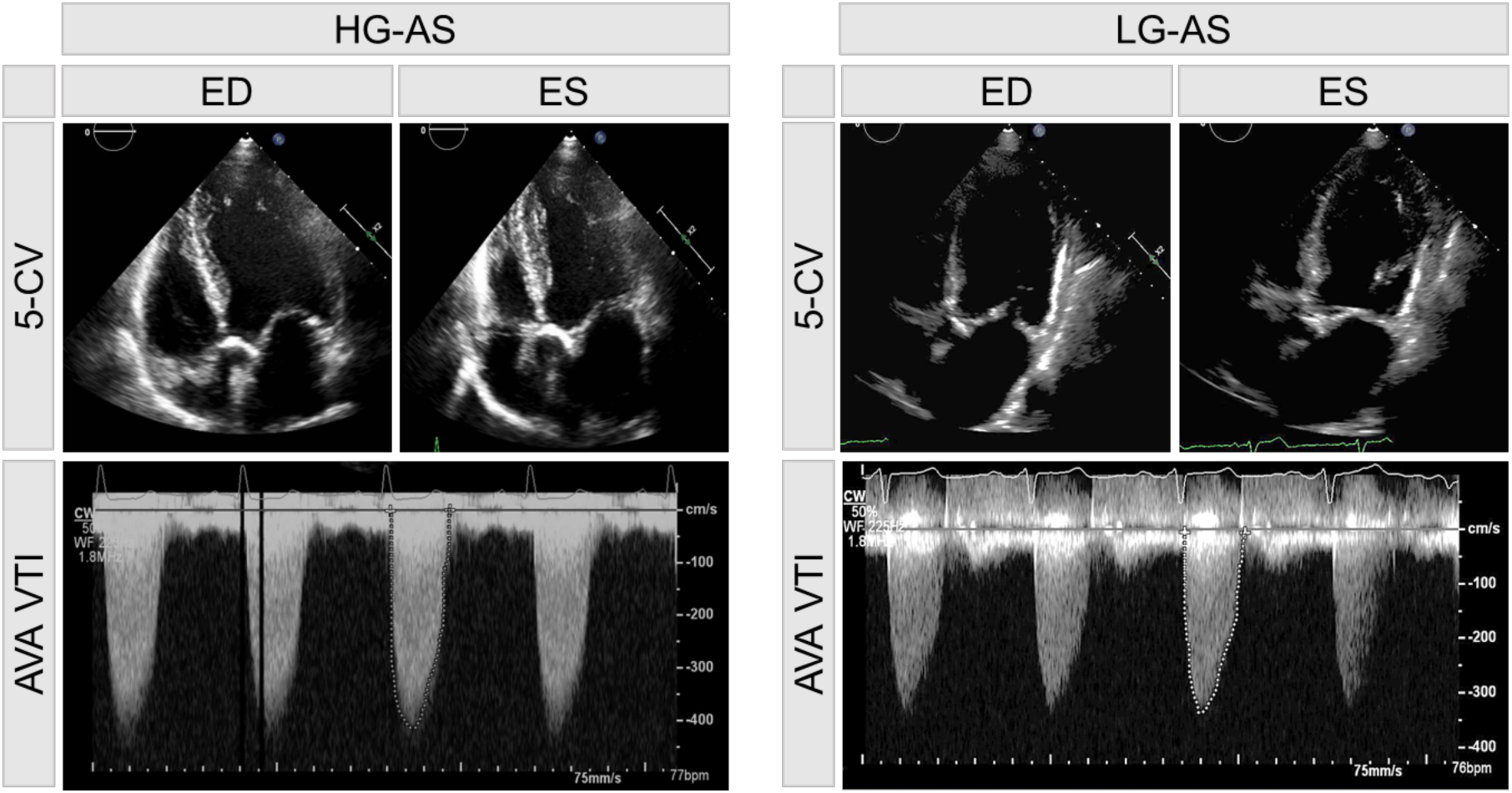
**Echocardiographic images of patient with high-gradient (HG) and low-gradient (LG) aortic stenosis (AS), respectively.** Pictures in the top showing a 5-chamber view (5-CV) in end-diastole (ED) and end-systole (ES). The corresponding derived aortic valve area velocity time integral (AVA VTI) signal is shown below.

LV-EF was calculated by Simpson’s biplane method using the manual tracing of the outline of the endocardial border on apical four and apical two-chamber view. To calculate relative wall thickness the formula (2 × posterior wall thickness/LVEDD) was used. Peak pulmonary artery systolic pressure was calculated using peak tricuspid regurgitation velocity (+ right atrial pressure). In order to validate echocardiographic findings of the study cohort, additional analyses of echocardiographic data obtained from a large clinical all-comer cohort were performed.

### Next generation sequencing

The alignment, normalization and analysis of the RNA-seq data were performed using the RStudio IDE with R version 4.2.2. The raw .fastq files were aligned to the GRCh38 genome using the align function in the Rsubread R package.[10] Only uniquely mapped reads were taken into account. The Rsubread function featureCounts was used to summarize aligned reads to features of the gene annotation file taken from GenCode (Release 40).[11] After adding a pseudocount of 1 to all features the expression profiles were normalized to GeTMM values.[12] The functions lmFit and eBayes from the limma package were used to check for differentially expressed genes. [13]

### Histology

After TAVI procedure, tissue samples were harvested from the basal anteroseptum in the left ventricle using biopsy forceps (Proflex-Bioptom 7F). These samples were then fixed in 10% paraformaldehyde and embedded in paraffin for further analysis. MF was evaluated in a blinded manner, using quantitative morphometry through Olympus Software cell-Sens 1.6. It was defined as the proportion of blue-stained area, indicating collagen presence, in sections of biopsy samples stained with Masson’s trichrome. This proportion was determined by comparing the blue-stained area to the total tissue area.

### Cardiovascular magnetic resonance imaging

#### CMR Imaging protocol

CMR imaging was performed on a 3.0-Tesla Magnetom Skyra MRI scanner (Siemens Healthcare, Erlangen, Germany) using a 32-channel cardiac surface receiver coil. Electrocardiography-gated balanced steady state free precession (b-SSFP) images of long-axis two, three- and four-chamber views (2-, 3-, 4-CV) as well as short-axis (SAX) stacks were acquired for functional myocardial assessments with the following typical image parameters: 25 frames per cardiac cycle, time of echo (TE) 1.5 ms, time of repetition (TR) 55 ms, flip angle 55°, 7 mm slice thickness with 7.7 mm inter-slice gap. Conventional 5(3)3 Modified Look-Locker Inversion Recovery (MOLLI) sequences (FOV of 360 x 306.6mm^2^, in-plane resolution 1.41 x 1.41 x 8mm^3^, TR 280ms, TE 1.12ms, TI 180ms, flip angle 35°, bandwidth 1085 Hz/pixel with total acquisition of 11 heart beats) for T1-mapping were performed ahead of admission of a gadolinium contrast bolus (0.15 mmol/kg bodyweight) and 20 minutes after. Phase-sensitive inversion-recovery-gradient echo sequences were acquired for late gadolinium enhancement (LGE) analyses 15-20 minutes after the gadolinium bolus injection with the following typical imaging parameters: TR 700 ms; TE 1.24 ms; flip angle 40°; slice thickness 7 mm and individually adjusted inversion times typically between 300 and 400 ms) (Figure 2).

**Figure 2:**
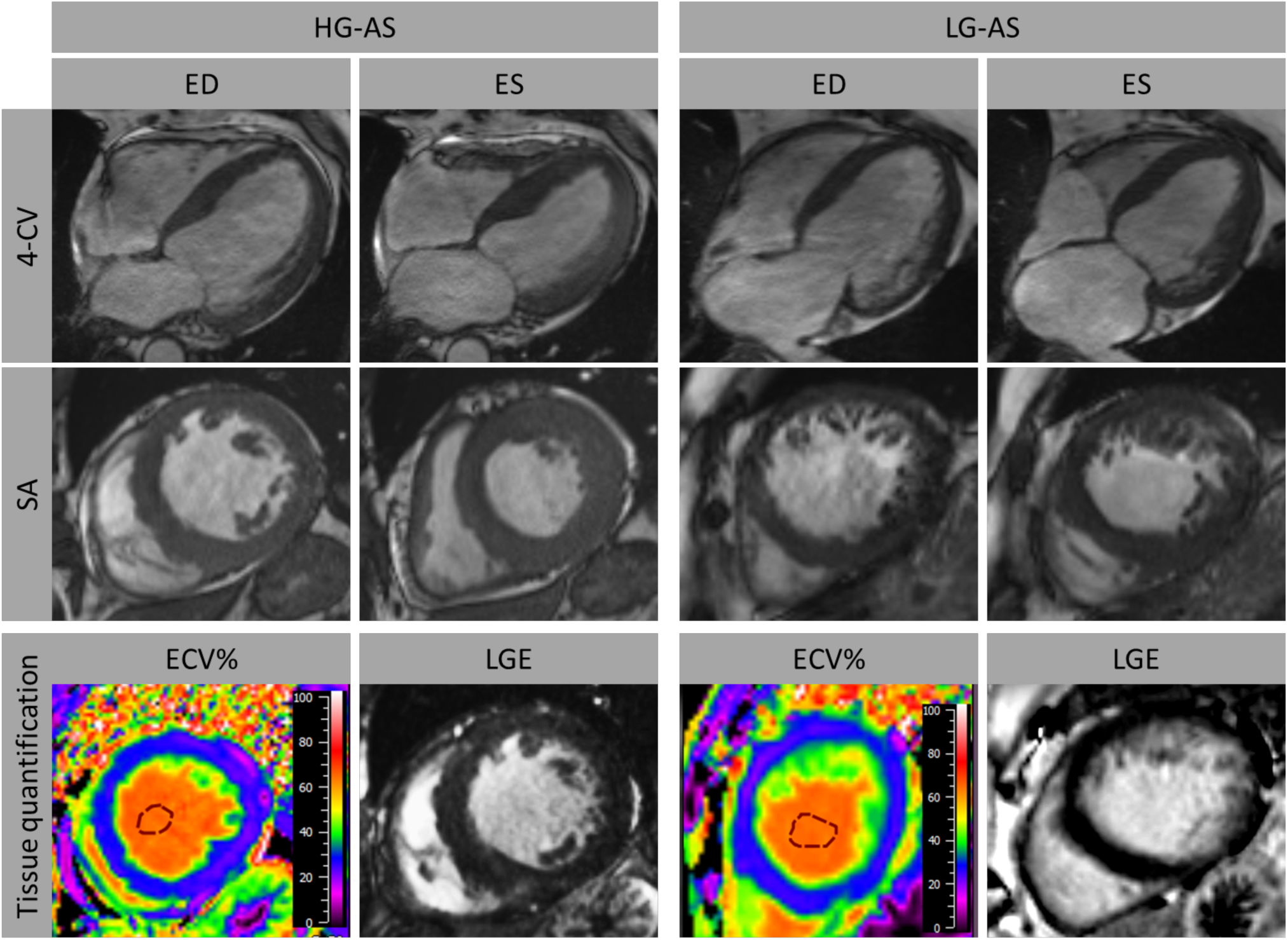
**Cardiovascular magnetic resonance imaging in patients with high-gradient (HG) and low-gradient (LG) aortic stenosis (AS).** Myocardial function was analyzed in long-axis orientation including 4-chamber view (CV) as well as short-axis (SA) orientation. Furthermore, tissue characterization comprising calculation of extracellular volume (ECV%) and late gadolinium enhancement (LGE) was performed. ED: end-diastole; ES: end-systole.

#### CMR Image Analysis

Based on b-SSFP images strain assessments were performed applying dedicated CMR-feature tracking (CMR-FT) evaluation software (Medis Medical Imaging Systems, Leiden, The Netherlands). Epi- and endocardial borders were manually delineated at end-diastole and - systole in 2-, 3- and 4-CV long axis orientations to analyze global longitudinal strain (GLS). Likewise, atrial endocardial contours were delineated in 2- and 4-CV images for the quantification of LA reservoir (LA Es), conduit (LA Ee) and booster pump strain (LA Ea) as described elsewhere.[14] For ventricular volumetric analyses epi- and endocardial borders were manually delineated in SAX stacks covering the entire LV. T1-mapping based assessment of extracellular volume (ECV%) reflecting myocardial tissue that is not occupied by cells was conducted based on motion-corrected MOLLI sequences. The definition of ECV% was as follows: ECV% = (1 – hematocrit) * [Δ R1 myocardium]/ [Δ R1 blood] according to guideline recommendations.[15] In all T1 weighted images regions of interest were drawn in excluding any LGE areas and careful visual reevaluation of the delineated regions was performed to avoid partial volume effects due to blood pool or adjacent non-myocardial structures. Using SAX images of inversion recovery sequences, LGE analyses were performed by defining a 3 standard deviations (SD) threshold of signal intensity for the detection of ischemic and non-ischemic LGE enhancement after manual epi- and endocardial border delineation as previously established.[16]

### Statistical analysis

Statistical analysis was performed in Graph Pad prism version 9.0. Continuous variables are presented as mean ± standard deviation. Categorial data were presented as frequency and percentage. Variables were tested for normal distribution by Shapiro-Wilk test. Subgroups were compared by two-sided T-test, one-way analysis of variance or Mann-Whitney-U test, if appropriate. For categorical variables, differences between HG- and LG-group were evaluated by Fisher exact test. A p-value of <0.05 was considered statistically significant.

## Results

### Baseline characteristics

98 patients with severe AS and impaired left ventricular systolic function, who were scheduled for TAVI at the University Medical Center Göttingen between January 2017 and December 2020 were included to this study (Figure 3). 40 patients were classified as HG-AS and 58 patients as LG-AS (pseudo stenosis was ruled out either by stress test or calcium score imaging). The mean age of the overall study population was 79.3 (± 6.8) years and participants were predominantly male (75%). Cardiovascular risk factors such as dyslipidemia, arterial hypertension, and diabetes mellitus were present in most of the patients (Table 1). On admission, majority of the patients showed signs of cardiac decompensation coming along with severe symptoms (NYHA III or IV). LG-AS patients showed no significant difference for chronic coronary syndrome (HG: 65% vs. LG: 77.6%; p=0.24), but more prior coronary interventions (HG: 17.5% vs. LG: 44.8%; p=0.005) or coronary artery bypass graft (CABG) (HG: 2.5% vs. LG: 22.4%; p=0.007). Furthermore, atrial fibrillation (AF) occurred more frequently in LG-AS patients (HG: 30% vs. LG: 51.7%; p=0.03), which also correlated with an increased rate of AF on admission (HG: 12.5% vs. 41%; p=0.0029) (Supplemental Table 1). Neither NT-proBNP (HG: 8707 ± 9109 ng/L vs. LG: 9460 ± 14228 ng/L; p=0.3) nor the 6MWT-distances differed significantly between the two subgroups. (HG: 197.9 ± 130.8 m vs. LG: 189.9 ± 118 m; p= 0.77). A more detailed overview of patient characteristics is presented in Table 1.

**Figure 3:**
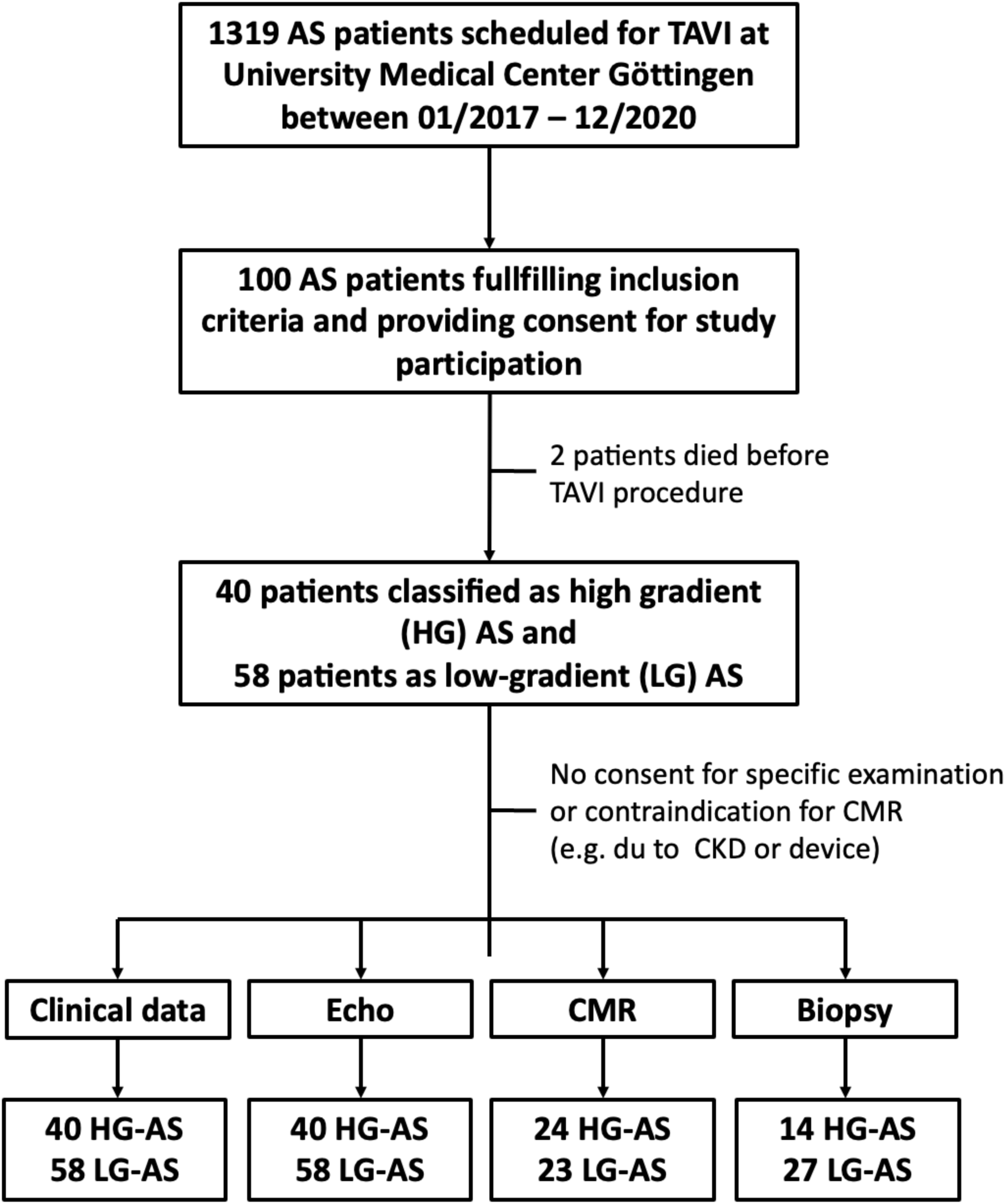
**Study flow chart**

**Table 1:**
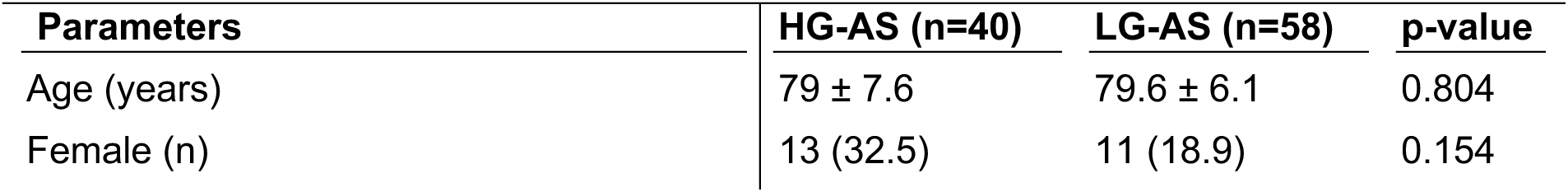
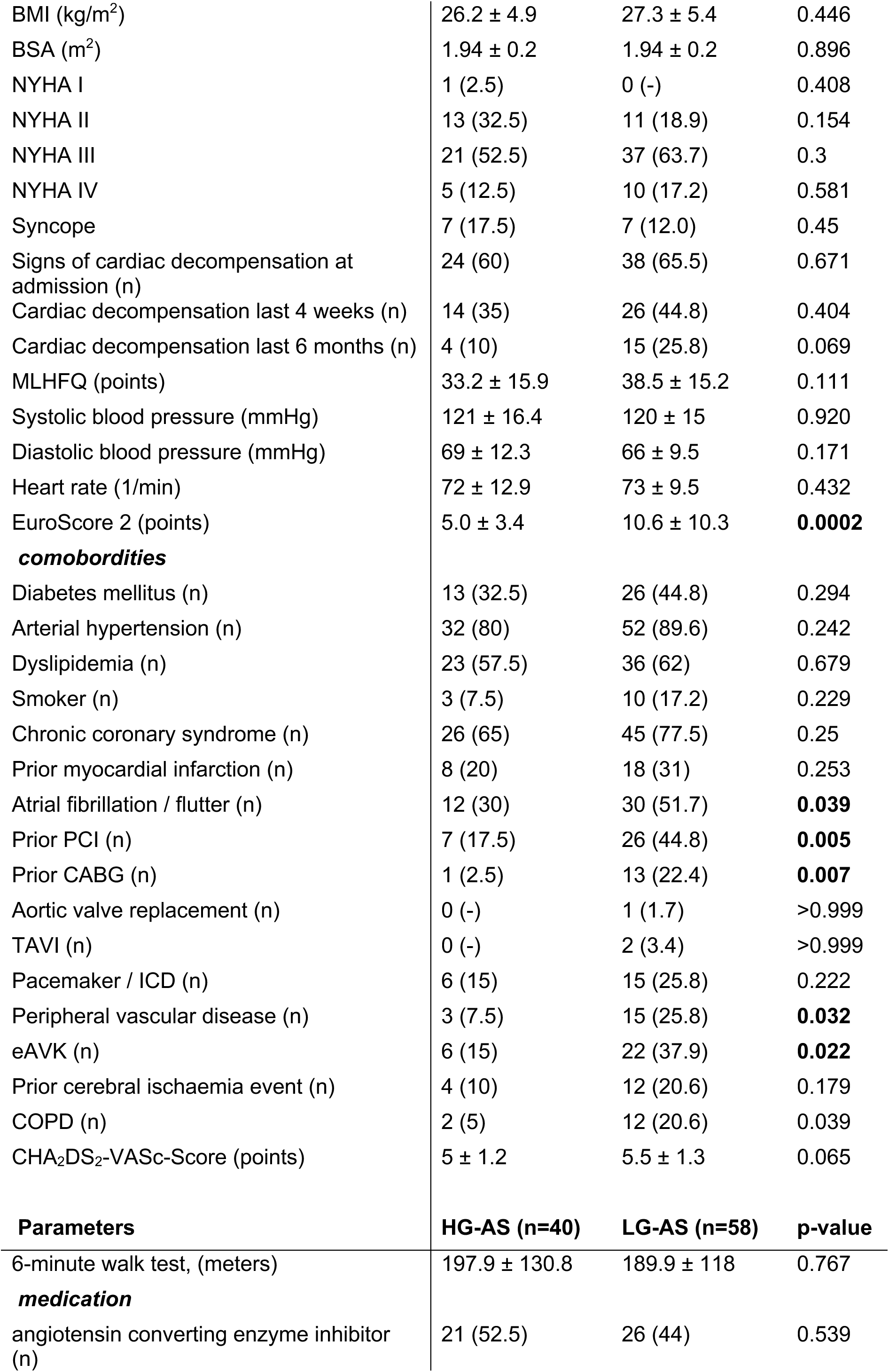
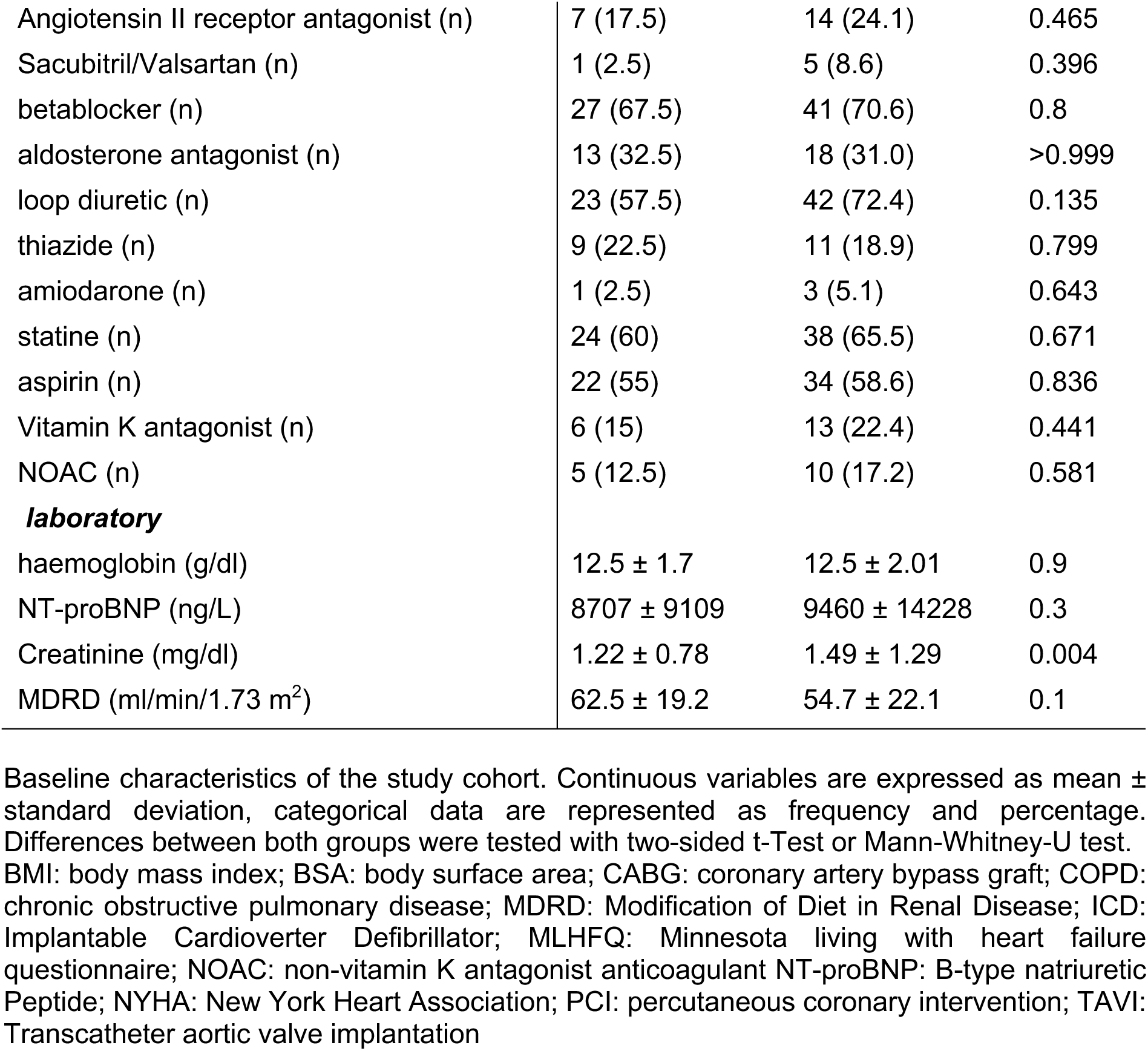
baseline clinical characteristics.

| Parameters | HG-AS (n=40) | LG-AS (n=58) | p-value |
| --- | --- | --- | --- |
| Age (years) | 79 ± 7.6 | 79.6 ± 6.1 | 0.804 |
| Female (n) | 13 (32.5) | 11 (18.9) | 0.154 |
| BMI (kg/m<sup>2</sup>) | 26.2 ± 4.9 | 27.3 ± 5.4 | 0.446 |
| BSA (m<sup>2</sup>) | 1.94 ± 0.2 | 1.94 ± 0.2 | 0.896 |
| NYHA I | 1 (2.5) | 0 (-) | 0.408 |
| NYHA II | 13 (32.5) | 11 (18.9) | 0.154 |
| NYHA III | 21 (52.5) | 37 (63.7) | 0.3 |
| NYHA IV | 5 (12.5) | 10 (17.2) | 0.581 |
| Syncope | 7 (17.5) | 7 (12.0) | 0.45 |
| Signs of cardiac decompensation at admission (n) | 24 (60) | 38 (65.5) | 0.671 |
| Cardiac decompensation last 4 weeks (n) | 14 (35) | 26 (44.8) | 0.404 |
| Cardiac decompensation last 6 months (n) | 4 (10) | 15 (25.8) | 0.069 |
| MLHFQ (points) | 33.2 ± 15.9 | 38.5 ± 15.2 | 0.111 |
| Systolic blood pressure (mmHg) | 121 ± 16.4 | 120 ± 15 | 0.920 |
| Diastolic blood pressure (mmHg) | 69 ± 12.3 | 66 ± 9.5 | 0.171 |
| Heart rate (1/min) | 72 ± 12.9 | 73 ± 9.5 | 0.432 |
| EuroScore 2 (points) | 5.0 ± 3.4 | 10.6 ± 10.3 | **0.0002** |
| ***comorbordities*** |  |  |  |
| Diabetes mellitus (n) | 13 (32.5) | 26 (44.8) | 0.294 |
| Arterial hypertension (n) | 32 (80) | 52 (89.6) | 0.242 |
| Dyslipidemia (n) | 23 (57.5) | 36 (62) | 0.679 |
| Smoker (n) | 3 (7.5) | 10 (17.2) | 0.229 |
| Chronic coronary syndrome (n) | 26 (65) | 45 (77.5) | 0.25 |
| Prior myocardial infarction (n) | 8 (20) | 18 (31) | 0.253 |
| Atrial fibrillation / flutter (n) | 12 (30) | 30 (51.7) | **0.039** |
| Prior PCI (n) | 7 (17.5) | 26 (44.8) | **0.005** |
| Prior CABG (n) | 1 (2.5) | 13 (22.4) | **0.007** |
| Aortic valve replacement (n) | 0 (-) | 1 (1.7) | >0.999 |
| TAVI (n) | 0 (-) | 2 (3.4) | >0.999 |
| Pacemaker / ICD (n) | 6 (15) | 15 (25.8) | 0.222 |
| Peripheral vascular disease (n) | 3 (7.5) | 15 (25.8) | **0.032** |
| eAVK (n) | 6 (15) | 22 (37.9) | **0.022** |
| Prior cerebral ischaemia event (n) | 4 (10) | 12 (20.6) | 0.179 |
| COPD (n) | 2 (5) | 12 (20.6) | 0.039 |
| CHA<sub>2</sub>DS<sub>2</sub>-VASc-Score (points) | 5 ± 1.2 | 5.5 ± 1.3 | 0.065 |
| **Parameters** | **HG-AS (n=40)** | **LG-AS (n=58)** | **p-value** |
| 6-minute walk test, (meters) | 197.9 ± 130.8 | 189.9 ± 118 | 0.767 |
| ***medication*** |  |  |  |
| angiotensin converting enzyme inhibitor (n) | 21 (52.5) | 26 (44) | 0.539 |
| Angiotensin II receptor antagonist (n) | 7 (17.5) | 14 (24.1) | 0.465 |
| Sacubitril/Valsartan (n) | 1 (2.5) | 5 (8.6) | 0.396 |
| betablocker (n) | 27 (67.5) | 41 (70.6) | 0.8 |
| aldosterone antagonist (n) | 13 (32.5) | 18 (31.0) | >0.999 |
| loop diuretic (n) | 23 (57.5) | 42 (72.4) | 0.135 |
| thiazide (n) | 9 (22.5) | 11 (18.9) | 0.799 |
| amiodarone (n) | 1 (2.5) | 3 (5.1) | 0.643 |
| statine (n) | 24 (60) | 38 (65.5) | 0.671 |
| aspirin (n) | 22 (55) | 34 (58.6) | 0.836 |
| Vitamin K antagonist (n) | 6 (15) | 13 (22.4) | 0.441 |
| NOAC (n) | 5 (12.5) | 10 (17.2) | 0.581 |
| ***laboratory*** |  |  |  |
| haemoglobin (g/dl) | 12.5 ± 1.7 | 12.5 ± 2.01 | 0.9 |
| NT-proBNP (ng/L) | 8707 ± 9109 | 9460 ± 14228 | 0.3 |
| Creatinine (mg/dl) | 1.22 ± 0.78 | 1.49 ± 1.29 | 0.004 |
| MDRD (ml/min/1.73 m<sup>2</sup>) | 62.5 ± 19.2 | 54.7 ± 22.1 | 0.1 |

Baseline characteristics of the study cohort. Continuous variables are expressed as mean ± standard deviation, categorical data are represented as frequency and percentage. Differences between both groups were tested with two-sided t-Test or Mann-Whitney-U test. BMI: body mass index; BSA: body surface area; CABG: coronary artery bypass graft; COPD: chronic obstructive pulmonary disease; MDRD: Modification of Diet in Renal Disease; ICD: Implantable Cardioverter Defibrillator; MLHFQ: Minnesota living with heart failure questionnaire; NOAC: non-vitamin K antagonist anticoagulant NT-proBNP: B-type natriuretic Peptide; NYHA: New York Heart Association; PCI: percutaneous coronary intervention; TAVI: Transcatheter aortic valve implantation

### Severity of AS and left ventricular geometry

Echocardiographic characteristics are presented in Table 2. AVA was slightly smaller in HG-patients compared to LG-patients. There was no difference in LVEF (HG: 37 ± 9.7% vs. LG: 33.9 ± 9.2%; p =0.1), LVEDD (HG: 51 ± 8.7 mm vs. LG: 54 ± 7.9 mm, p=0.12), relative wall thickness (HG: 0.54 ± 0.16 vs. LG: 0.48 ± 0.12, p=0.25) or LVMI (HG: 170 ± 36.5 g/m² BSA vs. LG: 166.8 ± 41.4 g/m² BSA; p=0.61). Similar results were found in CMR analyses of these patients (Table 3). These echocardiographic findings were consistent in the additionally analyzed all-comer cohort (Suppl. Table 2)

**Table 2:**
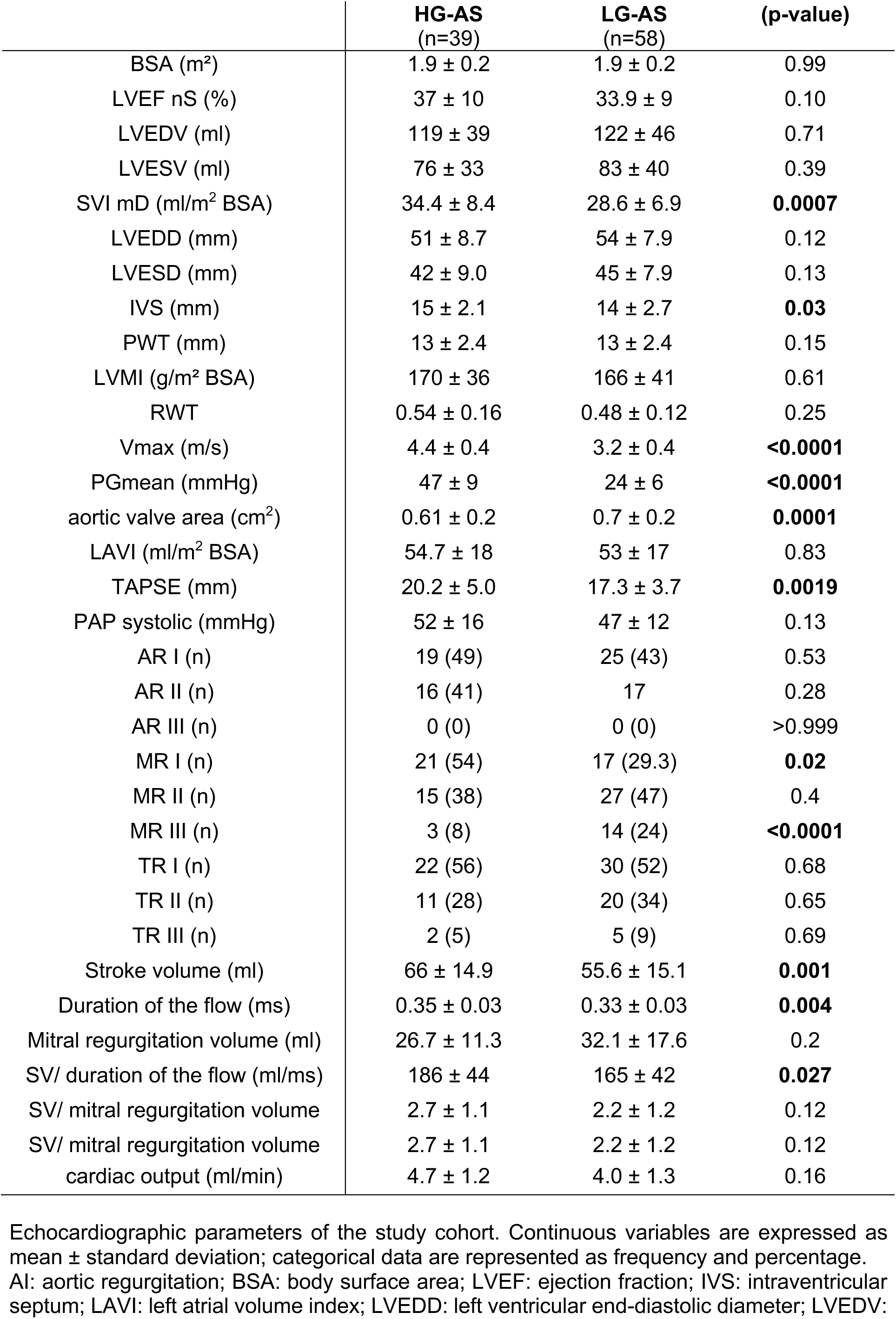
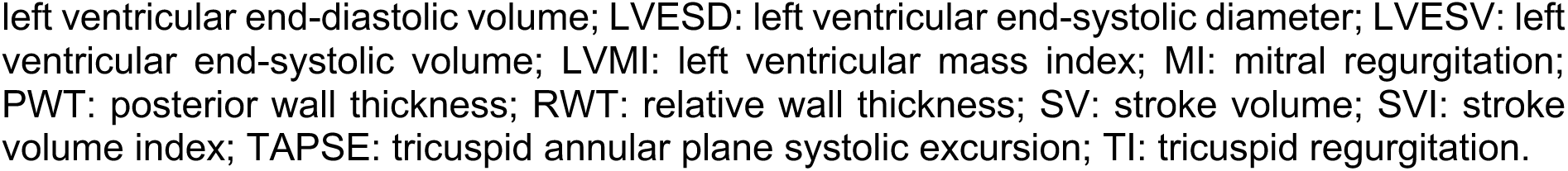
Echocardiographic characteristics.

**Table 3:** CMR characteristics.

| Parameter | HG-AS<br>(24) | LG-AS<br>(23) | p-value |
| --- | --- | --- | --- |
| niLGE volume (ml/ml<sup>2</sup>) | 8.9 (5.4-14.9) | 7.0 (2.7-13.7) | 0.31 |
| iLGE volume (ml/ml<sup>2</sup>) | 0.0 (0.0-4.0) | 0.0 (0.0-7.2) | 0.84 |
| Total LGE volume<br>(ml/ml<sup>2</sup>) | 13.2 (7.7-19.3) | 7.5 (4.9-21.3) | 0.35 |
| ECV% (%) | 25.8 (24.4-26.9) | 27.1 (26.2-29.6) | **0.012** |
| LV matrix volume<br>(ml/ml<sup>2</sup>) | 21.5 (18.0-26.3) | 22.1 (17.7-25.8) | 0.88 |
| LV cellular volume<br>(ml/ml<sup>2</sup>) | 65.2 (55.8-75.4) | 53.3 (46.0-68.1) | 0.16 |
| Total LVMV (ml/ml<sup>2</sup>) | 105.0 (90.1-117.8) | 90.5 (76.1-109.6) | 0.15 |
| LVEDVi (ml/ml<sup>2</sup>) | 101.5 (91.5-128.5) | 105.3 (80.9-132.9) | 0.8 |
| LVESVi (ml/ml<sup>2</sup>) | 58.1 (44.9-77.2) | 68.2 (40.7-90.1) | 0.87 |
| LVSVi (ml/ml<sup>2</sup>) | 50.4 (37.5-54.9) | 40.7 (34.7-48.7) | **0.03** |
| RV volume (ml/ml<sup>2</sup>) | 20.8 (16.7-29.8) | 22.2 (18.1-26.5) | 0.9 |
| RVEDVi (ml/ml<sup>2</sup>) | 72.5 (59.7-88.4) | 73.3 (58.5-89.4) | 0.98 |
| RVESVi (ml/ml<sup>2</sup>) | 35.6 (22.4-50.1) | 41.3 (31.7-56.5) | 0.31 |
| RVSVi (ml/ml<sup>2</sup>) | 36.7 (30.1-43.6) | 31.1 (24.7-35.8) | **0.045** |
| GLS (%) | -15.6 (-12.6- -17.5) | -13.4 (-9.7- -18.3) | 0.43 |
| GLS TTP (ms) | 373.4 (345.3-426.8) | 370.4 (352.5-400.0) | 0.83 |
| GCS (%) | -26.7 (-18.0- -34.6) | -20.9 (-14.8- -26.3) | 0.19 |
| GRS (%) | 40.1 (26.1-51.1) | 39.4 (31.4-50.4) | 0.87 |
| RV GLS (%) | -23.3 (-15.8- -29.7) | -23.0 (-15.5- -26.2) | 0.47 |
| LA Es (%) | 11.2 (7.8-16.6) | 8.7 (6.9-12.7) | 0.21 |
| LA Ee (%) | 4.4 (2.7-6.0) | 3.8 (1.3-6.6) | 0.67 |
| LA Ea (%) | 7.4 (4.5-9.4) | 5.4 (3.9-9.5) | 0.46 |
| RVEF (%) | 49.1 (38.1-62.4) | 43.7 (32.2-50.8) | **0.023** |
| LVEF (%) | 43.5 (37.3-53.1) | 38.0 (28.4-48.1) | 0.17 |
| RA Es (%) | 12.8 (10.1-17.4) | 12.7 (8.0-14.3) | 0.33 |
| RA Ee (%) | 6.1 (2.5-9.0) | 4.8 (2.0-5.9) | 0.14 |
| RA Ea (%) | 8.5 (5.8-11.5) | 7.0 (4.2-9.3) | 0.33 |
| CMR PCWP | 16.5 (14.5-21.1) | 19.0 (16.0-21.4) | 0.32 |

Cardiovascular magnetic resonance (CMR) imaging derived parameters. Variables are presented as median (interquartile range). Values of high-gradient (HG) and low-gradient (LG) aortic stenosis (AS) patients were compared using Mann-Whitney-U test. P-values in bold indicate a statistical significance.
CMR-derived characteristics of the study cohort; CMR PCWP: pulmonary capillary wedge pressure (CMR derived); ECV%: extracellular volume fraction; GCS: global circumferential strain; GLS: global longitudinal strain; GLS TTP: global longitudinal strain time to peak; GRS: global radial strain; iLGE: ischemic late-gadolinium-enhancement; LA Ea: left atrial booster pump strain; LA Ee: left atrial conduit strain; LA Es: left atrial reservoir strain; LGE: late-gadolinium-enhancement; LVEF: left ventricular ejection fraction; LVEDVi: left ventricular end-diastolic volume index; LVESVi: left ventricular end-systolic volume index; LVSVi: left ventricular stroke volume index; niLGE: non-ischemic late-gadolinium-enhancement; RA Ea: right atrial booster pump strain; RA Ee: right atrial conduit strain; RA Es: right atrial reservoir strain; RVEDVi: right ventricular end-diastolic volume index; RVEF: right ventricular ejection fraction; RV GLS: right ventricular global longitudinal strain; RVESVi: right ventricular end-systolic volume index;

**Table 4:** Echocardiographic characteristics classified according to LV-EF.

|  | **HG-AS,**<br>**EF >35%**<br>**(n=28)** | **HG-AS,**<br>**EF < 35%**<br>**(n=11)** | **LG-AS,**<br>**EF >35%**<br>**(n=29)** | **LG-AS,**<br>**EF <35%**<br>**(n=29)** | **p-value** |
| --- | --- | --- | --- | --- | --- |
| BSA (m<sup>2</sup>) | 1.9 ± 0.23 | 2.0 ± 0.24 | 1.9 ± 0.23 | 1.9 ± 0.21 | 0.17 |
| LVEF nS (%) | 42 ± 5 | 24 ± 6 **(a)** | 42 ± 4.0 | 26 ± 5 **(d)** | **< 0.001** |
| LVEDV (ml) | 106 ± 36 | 153 ± 32**(a)** | 100 ± 27 | 144 ± 50 **(d)** | **< 0.001** |
| LVESV (ml) | 61 ± 23.3 | 116 ± 26 **(a)** | 58 ± 18 | 107 ± 40**(d)** | **< 0.001** |
| SVI mD<br>(ml/m<sup>2</sup> BSA) | 36.8 ± 7.1 | 29.5 ± 5.8 **(a)** | 27.4 ± 6.4 | 29.7 ± 7.1 | **< 0.001** |
| LVEDD (mm) | 48.5 ± 7 | 59.8 ± 5 **(a)** | 51.2 ± 7 | 57.1 ± 7 **(d)** | **< 0.001** |
| LVESD (mm) | 38.7 ± 6 | 52.3 ± 6 **(a)** | 41.7 ± 7 | 48.6 ± 7 **(d)** | **< 0.001** |
| IVS (mm) | 16.1 ± 2.2 | 15.0 1.9 | 14.7 ± 3.1 | 14.5 ± 2.2 | 0.09 |
| PWT (mm) | 13.9 ± 2.0 | 12.4 ± 2.4 | 12.6 ± 2.2 | 12.9 ± 2.5 | 0.16 |
| LVMI (g/m<sup>2</sup><br>BSA) | 165 ± 35 | 189 ± 28 | 154 ± 33 | 179 ± 44 | **0.026** |
| Vmax (m/s) | 4.52 ± 0.33 | 4.2 ± 0.2 | 3.1 ± 0.38 **(b)** | 3.3 ± 0.46 **(c)** | **< 0.001** |
| Pmean<br>(mmHg) | 49 ± 8.3 | 44 ± 3.4 | 22.6 ± 5.89 **(b)** | 26.4 ± 6.8 **(c)** | **< 0.001** |
| aortic valve<br>area (cm<sup>2</sup>) | 0.63 ± 0.17 | 0.58 ± 0.10 | 0.7 ± 0.15 | 0.77 ± 0.16<br>**(c)** | **0.001** |
| LAVI (ml/m<sup>2</sup><br>BSA) | 52.3 ± 16.3 | 61.4 ± 22.5 | 51.1 ± 13.5 | 57.0 ± 19.9 | 0.34 |
| TAPSE (mm) | 21.1 ± 4.9 | 17.7 ± 4.3 | 17.5 ± 4.2 **(c)** | 17.0 ± 3.0 | **0.032** |
| MI III | 3 (10) | 0 | 7 (29) | 7 (29) | 0.47 |
| PAP systolic<br>(mmHg) | 49 ± 14.9 | 62 ± 14.4 | 47 ± 10.6 | 48 ± 14.2 | 0.055 |
| regurgitation<br>volume (mitral)<br>(ml) | 28.1 ± 11.9 | 22.4 ± 7.9 | 32.7 ± 15.3 | 31.7 ± 19.1 | 0.6 |
| stroke volume<br>(ml) | 67 ± 14 | 62 ± 15.1 | 53 ± 15.8 | 58 ± 13 | **0.006** |
| duration of the<br>flow (ms) | 0.36 ± 0.03 | 0.36 ± 0.03 | 0.33 ± 0.03 | 0.33 ± 0.03 | **0.043** |
| SV/ duration of<br>the flow<br>(ml/ms) | 190 ± 43 | 174 ± 45.1 | 158 ± 43.6 | 174 ± 39 | 0.053 |
| SV/ mitral<br>regurgitation<br>volume | 2.7 ± 1.1 | 2.5 ± 0.8 | 2.0 ± 1.0 | 2.4 1.1 | 0.38 |
| cardiac output | 3.9 ± 1.2 | 4.9 ± 1.7 | 3.9 ± 1.2 | 4.2 ± 1.2 | 0.07 |

a) HG >35 % vs. HG < 35 %; b) HG >35 % vs. LG > 35 %; c) HG <35 % vs. LG < 35 %; d) LG > 35 % vs. LG < 35 %
Echocardiographic parameter, classified according to LV-EF above and below 35%. Continuous variables are expressed as mean ± standard deviation. categorical data are represented as frequency and percentage. AI: aortic insufficiency; BSA: body surface area; LVEF: left-ventricular ejection fraction; IVS: intraventricular septum; LAVI: left atrial volume index; LVEDD: left ventricular end-diastolic diameter; LVEDV: left ventricular end-diastolic volume; LVESD: left ventricular end-systolic diameter; LVESV: left ventricular end-systolic volume; LVMI: left ventricular mass index; MI: mitral insufficiency; PWT: posterior wall thickness; RWT: relative wall thickness; SV: stroke volume; SVI: stroke volume index; TAPSE: tricuspid annular plane systolic excursion; TI: tricuspid insufficiency.

The lower stroke volume (HG: 66 ± 14.9 ml vs. LG: 55.6 ± 15.1 ml; p=0.001) in LG-AS patients, however, could be explained by a shorter flow duration via the stenosed aortic valve (HG: 0.35 vs. LG: 0.33; p=0.004). This indicates that despite comparable LVEF and LVEDD the left ventricle cannot generate the same pressure time integral. Characterization of the LV by CMR did not show a difference in left ventricular strain or volumes, while ECV% was significantly higher in LG-AS compared to HG-AS patients (27.1 % [26.2-29.6] vs. 25.8 [24.4-26.9], p=0.012) (Table 3).

Notably, a higher prevalence of severe mitral regurgitation (HG: 8% vs. LG: 24%; p<0.001) and an impaired right ventricular dysfunction (RVSVi: 36.7 vs. 31.1 ml/m2; p=0.045; TAPSE: 20.2 vs. 17.3mm, p=0.002) were detected in the LG-group. Furthermore, impaired hemodynamics especially affecting the right heart might be additionally indicated by regional differences in LV strain. Comparing septal vs. lateral GLS within each group revealed a significantly reduced septal GLS in the LG group (septal GLS −8.2 % vs. lateral GLS −12.7 %, p=0.008), which might be caused by raised loading pressures in the right ventricle hampering septal contractility (Table 5). In the HG group there was no difference between septal and lateral GLS (septal GLS −11.4 % vs. lateral GLS −12.0 %, p=0.22)

**Table 5:** Comparison of septal and lateral global longitudinal strain.

|  | **Septal** | **Lateral** | **p-value** |
| --- | --- | --- | --- |
| **HG-AS GLS** | -11.4 % | -12.0 % | 0.22 |
| **HG-AS GLS TTP** | 397 ms | 385 ms | **0.03** |
| **LG-AS GLS** | -8.2 % | -12.7 % | **0.008** |
| **LG-AS GLS TTP** | 366 ms | 369 ms | 0.49 |

|  | **HG-AS** | **LG-AS** | **p-value** |
| --- | --- | --- | --- |
| **Septal GLS** | -11.4 % | -8.2 % | 0.32 |
| **Septal GLS TTP** | 397 ms | 366 ms | 0.27 |
| **Lateral GLS** | -12.0 % | -12.7 % | 0.63 |
| **Lateral GLS TTP** | 385 ms | 369 ms | 0.33 |

AS: aortic stenosis, HG: high-gradient, GLS: global longitudinal strain, LG: low-gradient, TTP: time to peak.

Histology or next-generation sequencing did not show a difference in fibrosis nor global gene expression profile (Figure 4) between HG- and LG-AS patients. Furthermore, neither PITP-levels (HG: 151 ± 33 % vs. LG: 144 ± 40 %; p= 0.58), nor CITP (HG: 10.8 ± 9.5 % vs. LG: 12.3 ± 11.9 %; p= 0.65) and CIPT/MMP1-ratio differed significantly (HG: 2.6 ± 2.2 % vs. LG: 3.2 ± 3.1 %; p= 0.46). Furthermore, CMR-derived PCWP did not differ between both groups (16.5 [14.5-21.1] vs. 19.0 [16.0-21.4], p=0.32) suggesting no difference in preload induced contractile activation according to the Frank-Starling-mechanism.

**Figure 4:**
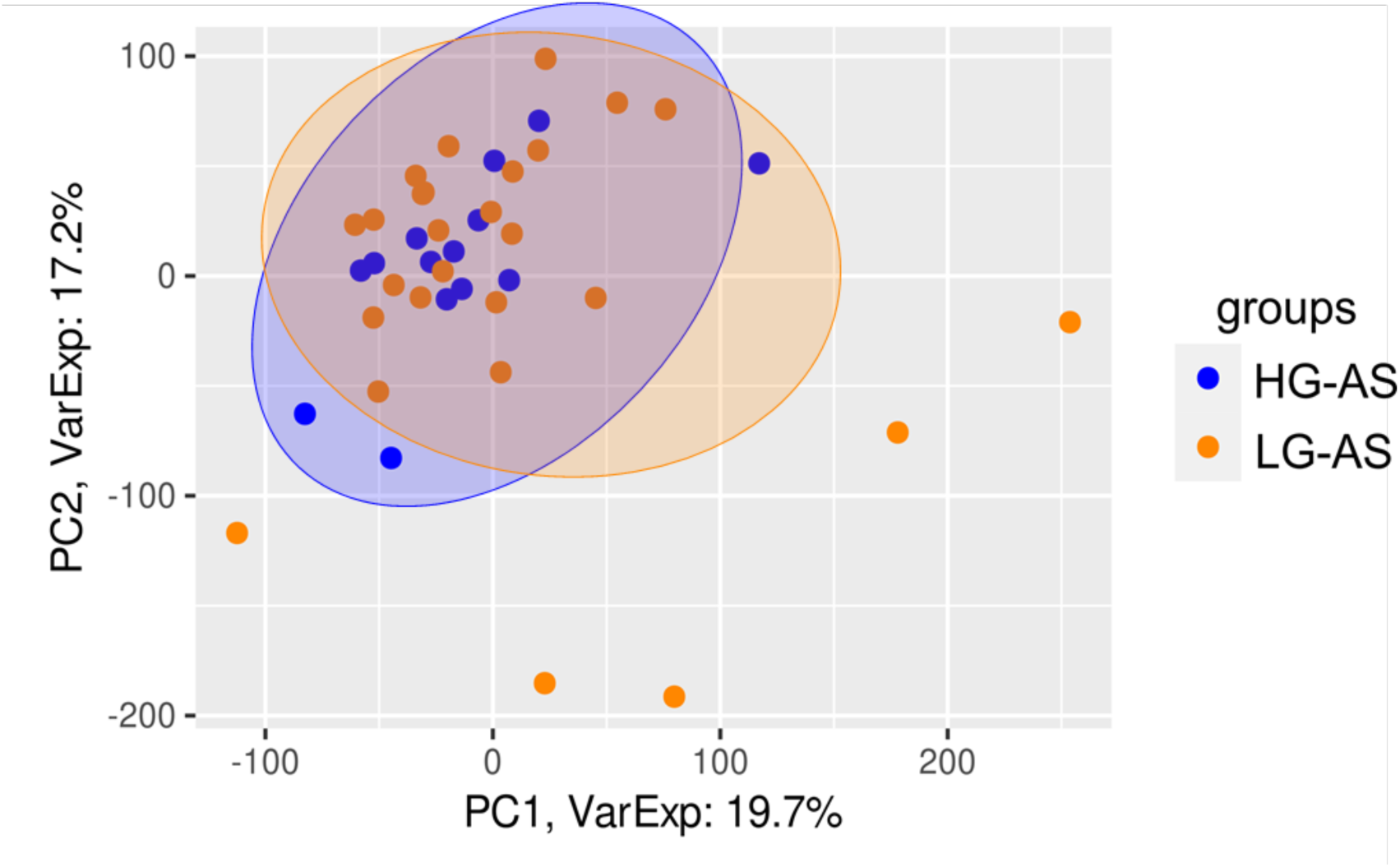
Principal component analysis (PCA) was employed to depict single LV biopsy samples obtained from individual patients. Each circle within the plot represents the projection of data from individual patient samples onto components 1 and 2, along with the percentage of total variance (x/y axis legend). Each patient’s dataset is represented by two technical replicates, and any outlier replicates have been excluded (14 HG-AS and 27 LG-AS)

## Discussion

This study comprehensively investigated characteristics of severe HG- and LG-AS patients with reduced LVEF and there are some major findings to be considered:

1. There were no significant differences in left ventricular function and volumetric analyses.
2. No differences between subgroups were detected via histological, gene expression and circulating biomarker-based assessment.
3. Right ventricular function was significantly impaired in LG-AS patients compared to HG-AS.
4. Prevalence of atrial fibrillation and high-grade mitral regurgitation was significantly higher in LG-AS patients.

To compensate increased afterload caused by severe AS, the myocardium hypertrophes with subsequent remodeling processes leading to increased myocardial fibrosis. As the disease progresses, compensatory capabilities of the myocardium begin to fail resulting in a decrease of myocardial function.^7,^[17]^,23^ Importantly, patients with severe AS and reduced LVEF have worse outcome than AS patients with preserved LVEF.[18] Nevertheless, there are two different subgroups of AS patients with reduced LVEF – one subgroup generating high transaortic pressure gradients, whereas the other group has only low pressure gradients, suggesting different left ventricular responses on pressure overload. However, there were no significant differences neither in LV function comprising strain and volumetry, nor in histological and NGS assessment between both subgroups. The only LV parameter that significantly differed was CMR-derived ECV%, which is considered as a surrogate parameter for diffuse myocardial fibrosis. Since this is in contrast to the histological and NGS findings, it remains speculative, whether these results are caused by the limitation of a local biopsy, whereas ECV% is more sensitive to detect global remodeling processes. [19] It is noteworthy, that AS is known to exhibit a characteristic diffuse myocardial fibrosis pattern.[16,20] Therefore, when evaluating myocardial fibrosis using methods like biopsy or CMR, it’s crucial to account for factors like location, sampling methods, and technical analysis aspects. Invasive biopsies are limited by size and sampling precision issues, whereas LGE and ECV measurements encompass different myocardial regions, providing complementary information, which should be considered when interpreting the (combination of) different results and parameters.

In contrast, there was a significant reduction in right ventricular function (both in echocardiography and CMR) in the LG-AS subgroup. Following considerations of the Frank-Starling mechanism, RV dysfunction typically develops as a consequence of pressure and/or volume overload. This might be caused by an impaired left heart performance, chronic lung disease or pulmonary hypertension.[21] In addition, the occurrence of LV hypertrophy/LV dysfunction was shown to influence RV function by a pathological downstream response to pressure overload. In this context, in this study documented impaired septal GLS contraction in LG-AS patients might further explain deterioration of RV function.

On the one hand, the current results do not show relevant differences in left heart (dys-)function and consequently one might speculate, whether the deterioration of RVEF is rather caused by an intrinsic RV myopathy than as a mere consequence of LV failure. In line with these considerations, the CMR-derived PCWP did not differ between both subgroups and, therefore, potentially increased LV filling pressures might not explain an impaired RVEF and rather points towards an intrinsic right heart pathology. However, especially the right ventricular and atrial strain values were not reduced in the LG-AS subgroup. Since deformation parameters like CMR-derived strain have been demonstrated to be even more sensitive parameters to detect functional deterioration than volumetric alterations and that functional changes precede myocardial geometric alterations, a fundamental RV contractile deterioration cannot be demonstrated.[22,23] Therefore, a decreased RVEF might rather be caused by impaired (LV and RV) hemodynamics than by a functional deterioration per se.

On the other hand, further explanations for altered hemodynamics and a lower transaortic gradient might be comorbidities like AF or a higher prevalence of mitral regurgitation, which may result in an incapability of the left ventricle to generate a higher gradient across the stenotic aortic valve. Since the severity of functional MR in AS patients is often directly influenced by elevated systolic ventricular pressures, this might indicate at least slight differences in ventricular pressures/ hemodynamics that are not measurable by the applicated (non-invasive) parameters. [24,25]

Importantly, despite pathomechanistical considerations for different gradients, several studies have demonstrated substantial prognostic value both of right ventricular function and comorbidities in AS patients.[26,27] It is noteworthy that RVD has been demonstrated to be independently associated with adverse outcome after adjusting for other cardiac comorbidities.[26] However, an additional prognostic impact of cardiac comorbidities, such as AF or MR, beyond aortic stenosis and its subsequent impairment of myocardial performance has been shown in several studies. [28] Furthermore, comorbidities like AF have been linked with lower transaortic gradients and with adverse outcome in LG-AS patients, underlining its crucial influence on hemodynamics and prognosis in these patient cohorts. [29]

Regarding clinical implications, although guidelines and/or risk calculators include LV function or transaortic gradients for risk stratification and therapeutic decision making in AS patients, they disregard other prognostically important parameters. For example, RV function is not implemented in the EuroSCORE II and STS score despite its reported association with adverse outcome in patients undergoing TAVI. [30]

Hence, by comprehensive analyses of myocardial performance including CMR-derived right heart function as well as coexisting comorbidities, optimized timing of valve replacement and post-procedural monitoring could improve future management of AS patients undergoing TAVI. Further studies are needed to validate these findings and to transfer them into clinical practice.

## Limitations

Data were prospectively collected in our TAVI registry, without excluding comorbidities such as CAD, which makes the study population an all-comer cohort. However, the data were collected monocentrically and retrospectively queried. Furthermore, the global NGS data set showed no distinct difference when comparing the two subgroups, but we cannot exclude the possibility of significant differences within individual genes.

## Conclusion

In severe AS with reduced LVEF, patients with low transaortal gradient do not differ in left ventricular function or tissue composition compared to patients with high transaortal gradients. In contrast, right ventricular function is more restricted in the low gradient subgroup and additional cardiac comorbidities like atrial fibrillation or mitral regurgitation are more frequent than in patients with high transaortic gradients. Possessing additional important prognostic information, future diagnostic algorithms and treatment recommendations might therefore incorporate comprehensive assessment of right heart function and comorbidities for optimized treatment and patient management.

## Author contributions

SG & TL: conceptualization, data curation, formal analysis, investigation, methodology, writing—original draft; BEB: data acquisition; ME: methodology (NGS), writing—original draft; NP: formal investigation, methodology; MS: formal analysis, data curation, writing – original draft; JD: supervision, writing; EZ: data curation; MP: data acquisition, data curation, supervision; GH: resources, supervision, writing - original draft; AS: conceptualization, project administration resources, supervision, validation; KT: conceptualization, formal analysis, investigation, methodology, project administration, resources, supervision, validation, writing - original draft. All authors read and approved the final manuscript.

## Funding

The study was funded by the German Research Foundation (DFG, SFB 1002 D04).

## Data availability statement

Data are available from the corresponding author upon reasonable request.

## Acknowledgments

We especially thank the study nurses Swetlana Hartmann and Kristina Schröder for study organization.

## Declarations

The study was approved by the Ethics Committee of the University Medical Center Göttingen (Ethic committee Göttingen: 22/4/11) and complied with the Declaration of Helsinki. All individuals gave written informed consent before participating in the study.

## Abbreviations

AF: atrial fibrillation
AS: Aortic stenosis
AVR: Aortic valve replacement
CITP: Carboxy-terminal telopeptide of collagen type I
CITP:MMP1: Ratio of serum carboxy-terminal telopeptide of collagen type I to serum matrix metalloproteinase-1
CMR: Cardiovascular magnetic resonance imaging
ECV: Extracellular volumen
HG: High gradient
LG: Low gradient
LGE: Late gadolinium enhancement
LVEDD: Left ventricular end-diastolic diameter
LVEF: left-ventricular ejection fraction
MF: Myocardial fibrosis
MMP1: matrix metalloproteinase-1
MR: mitral regurgitation
NGS: next-generation-sequencing
NT-proBNP: N-terminal pro-brain natriuretic peptide
NYHA: New York Heart Association
PICP: C-terminal propeptide of procollagen type I
Pw: Pulsed wave
RVEF: Right ventricular ejection fraction
TAPSE: Tricuspid annular plane systolic excursion
TAVI: Transcatheter aortic valve implantation

---

Supplemental Table 1 – baseline electrocardiographic characteristics

Supplemental Table 2 – Echocardiographic parameters assessed by transthoracic echocardiography in an all comer clinical cohort

## Notes

### Competing Interest Statement

The authors have declared no competing interest.

### Funding Statement

German Research Foundation (DFG, CRC 1002, D1)

### Author Declarations

Ethics committee/IRB of university medical school goettingen gave ethical approval for this work.

### Summary of Updates

Author team.

## References

1 Iung B, Vahanian A. Epidemiology of valvular heart disease in the adult. Nat Rev Cardiol. 2011;8:162–72.

2 Iung B, Baron G, Butchart EG, et al. A prospective survey of patients with valvular heart disease in Europe: The Euro Heart Survey on Valvular Heart Disease. Eur Heart J. 2003;24:1231–43.

3 Bevan GH, Zidar DA, Josephson RA, et al. Mortality Due to Aortic Stenosis in the United States, 2008-2017. JAMA. 2019;321:2236–8.

4 Vahanian A, Beyersdorf F, Praz F, et al. 2021 ESC/EACTS Guidelines for the management of valvular heart disease: Developed by the Task Force for the management of valvular heart disease of the European Society of Cardiology (ESC) and the European Association for Cardio-Thoracic Surgery (EACTS). European Heart Journal. 2022;43:561–632.

5 Puls M, Beuthner BE, Topci R, et al. Impact of myocardial fibrosis on left ventricular remodelling, recovery, and outcome after transcatheter aortic valve implantation in different haemodynamic subtypes of severe aortic stenosis. European Heart Journal. 2020;41:1903–14.

6 Baron SJ, Arnold SV, Herrmann HC, et al. Impact of Ejection Fraction and Aortic Valve Gradient on Outcomes of Transcatheter Aortic Valve Replacement. J Am Coll Cardiol. 2016;67:2349–58.

7 Le Ven F, Freeman M, Webb J, et al. Impact of Low Flow on the Outcome of High-Risk Patients Undergoing Transcatheter Aortic Valve Replacement. Journal of the American College of Cardiology. 2013;62:782–8.

8 López B, Ravassa S, González A, et al. Myocardial Collagen Cross-Linking Is Associated With Heart Failure Hospitalization in Patients With Hypertensive Heart Failure. J Am Coll Cardiol. 2016;67:251–60.

9 Lang RM, Badano LP, Mor-Avi V, et al. Recommendations for cardiac chamber quantification by echocardiography in adults: an update from the American Society of Echocardiography and the European Association of Cardiovascular Imaging. J Am Soc Echocardiogr. 2015;28:1–39.e14.

10 Liao Y, Smyth GK, Shi W. The R package Rsubread is easier, faster, cheaper and better for alignment and quantification of RNA sequencing reads. Nucleic Acids Res. 2019;47:e47.

11 Frankish A, Diekhans M, Ferreira A-M, et al. GENCODE reference annotation for the human and mouse genomes. Nucleic Acids Research. 2019;47:D766–73.

12 Smid M, Coebergh van den Braak RRJ, van de Werken HJG, et al. Gene length corrected trimmed mean of M-values (GeTMM) processing of RNA-seq data performs similarly in intersample analyses while improving intrasample comparisons. BMC Bioinformatics. 2018;19:236.

13 Ritchie ME, Phipson B, Wu D, et al. limma powers differential expression analyses for RNA-sequencing and microarray studies. Nucleic Acids Research. 2015;43:e47.

14 Lange T, Schuster A. Quantification of Myocardial Deformation Applying CMR-Feature-Tracking—All About the Left Ventricle? Curr Heart Fail Rep. 2021;18:225–39.

15 Messroghli DR, Moon JC, Ferreira VM, et al. Clinical recommendations for cardiovascular magnetic resonance mapping of T1, T2, T2* and extracellular volume: A consensus statement by the Society for Cardiovascular Magnetic Resonance (SCMR) endorsed by the European Association for Cardiovascular Imaging (EACVI). Journal of Cardiovascular Magnetic Resonance. 2017;19:75.

16 Treibel TA, López B, González A, et al. Reappraising myocardial fibrosis in severe aortic stenosis: an invasive and non-invasive study in 133 patients. European Heart Journal. 2018;39:699–709.

17 Hein S. Progression From Compensated Hypertrophy to Failure in the Pressure-Overloaded Human Heart: Structural Deterioration and Compensatory Mechanisms. Circulation. 2003;107:984–91.

18 Ito S, Miranda WR, Nkomo VT, et al. Reduced Left Ventricular Ejection Fraction in Patients With Aortic Stenosis. Journal of the American College of Cardiology. 2018;71:1313–21.

19 Schulz-Menger J, Wassmuth R, Abdel-Aty H, et al. Patterns of myocardial inflammation and scarring in sarcoidosis as assessed by cardiovascular magnetic resonance. Heart. 2006;92:399–400.

20 Lange T, Backhaus SJ, Beuthner BE, et al. Functional and structural reverse myocardial remodeling following transcatheter aortic valve replacement: a prospective cardiovascular magnetic resonance study. Journal of Cardiovascular Magnetic Resonance. 2022;24:45.

21 Eleid MF. Right Ventricular Function in TAVR: The Right Hand Knows What the Left Hand Is Doing∗. JACC: Cardiovascular Imaging. 2019;12:588–90.

22 Kim J, Yum B, Palumbo MC, et al. Left Atrial Strain Impairment Precedes Geometric Remodeling as a Marker of Post-Myocardial Infarction Diastolic Dysfunction. JACC: Cardiovascular Imaging. 2020;13:2099–113.

23 DeVore AD, McNulty S, Alenezi F, et al. Impaired left ventricular global longitudinal strain in patients with heart failure with preserved ejection fraction: insights from the RELAX trial. Eur J Heart Fail. 2017;19:893–900.

24 O’Sullivan CJ, Stortecky S, Bütikofer A, et al. Impact of mitral regurgitation on clinical outcomes of patients with low-ejection fraction, low-gradient severe aortic stenosis undergoing transcatheter aortic valve implantation. Circ Cardiovasc Interv. 2015;8:e001895.

25 Freitas-Ferraz AB, Lerakis S, Barbosa Ribeiro H, et al. Mitral Regurgitation in Low-Flow, Low-Gradient Aortic Stenosis Patients Undergoing TAVR: Insights From the TOPAS-TAVI Registry. JACC Cardiovasc Interv. 2020;13:567–79.

26 Asami M, Stortecky S, Praz F, et al. Prognostic Value of Right Ventricular Dysfunction on Clinical Outcomes After Transcatheter Aortic Valve Replacement. JACC Cardiovasc Imaging. 2019;12:577–87.

27 Bohbot Y, Guignant P, Rusinaru D, et al. Impact of Right Ventricular Systolic Dysfunction on Outcome in Aortic Stenosis. Circulation: Cardiovascular Imaging. 2020;13:e009802.

28 Tarantini G, Mojoli M, Windecker S, et al. Prevalence and Impact of Atrial Fibrillation in Patients With Severe Aortic Stenosis Undergoing Transcatheter Aortic Valve Replacement: An Analysis From the SOURCE XT Prospective Multicenter Registry. JACC: Cardiovascular Interventions. 2016;9:937–46.

29 Kolluri N, Oguz D, Scott CG, et al. Impact of atrial fibrillation in clinical outcomes of low gradient aortic stenosis. European Heart Journal. 2022;43:ehac544.1617.

30 Koschutnik M, Dannenberg V, Nitsche C, et al. Right ventricular function and outcome in patients undergoing transcatheter aortic valve replacement. Eur Heart J Cardiovasc Imaging. 2021;22:1295–303.

